# Predicting the unpredictable: how dynamic COVID-19 policies and restrictions challenge model forecasts

**DOI:** 10.1101/2021.09.30.21264273

**Authors:** Farah Houdroge, Anna Palmer, Dominic Delport, Tom Walsh, Sherrie L Kelly, Samuel W Hainsworth, Romesh Abeysuriya, Robyn M Stuart, Cliff C Kerr, Paul Coplan, David P Wilson, Nick Scott

**Author notes:** **Corresponding Author:** Farah Houdroge, PhD, Research Assistant, Mathematical Modeler, Modelling and Biostatistics Group, Burnet Institute, 85 Commercial Road, Melbourne, VIC, 3004, Australia.

## Abstract

**Introduction:** To retrospectively assess the accuracy of a mathematical modelling study that projected the rate of COVID-19 diagnoses for 72 locations worldwide in 2021, and to identify predictors of model accuracy.

**Methods:** Between June and August 2020, an agent-based model was used to project rates of COVID-19 infection incidence and cases diagnosed as positive from 15 September to 31 October 2020 for 72 geographic settings. Five scenarios were modelled: a baseline scenario where no future changes were made to existing restrictions, and four scenarios representing small or moderate changes in restrictions at two intervals. Post hoc, upper and lower bounds for number of diagnosed Covid-19 cases were compared with actual data collected during the prediction window. A regression analysis with 17 covariates was performed to determine correlates of accurate projections.

**Results:** The actual data fell within the lower and upper bounds in 27 settings and out of bounds in 45 settings. The only statistically significant predictor of actual data within the predicted bounds was correct assumptions about future policy changes (OR = 15.04; 95%CI 2.20-208.70; p=0.016).

**Conclusions:** For this study, the accuracy of COVID-19 model projections was dependent on whether assumptions about future policies are correct. Frequent changes in restrictions implemented by governments, which the modelling team was not always able to predict, in part explains why the majority of model projections were inaccurate compared with actual outcomes and supports revision of projections when policies are changed as well as the importance of policy experts collaborating on modelling projects.

## 1. Introduction

According to the World Health Organization (WHO), as of September 22, 2021, there have been more than 229 million confirmed cases of COVID-19 globally, causing the loss of more than 4.7 million lives [1].The wide and rapid spread of the SARS-CoV-2 virus precipitated a global health and economic crisis: the WHO reported a disruption to essential health services that affected 90% of countries between March and June 2020 [2], and the World Bank estimates that the global economy has experienced the deepest recession since the Second World War and that an upsurge in extreme poverty is to be expected [3, 4]

Governments have introduced physical distancing policies, hygiene protocols, and total or partial lockdowns to control virus transmission, but many countries who eased lockdown restrictions saw a resurgence of COVID-19 cases, making the development of a vaccine to combat COVID-19 one of the most urgent public health endeavors in modern history. As of September 22, 2021, there were one vaccine approved and three approved for emergency use in the United States, four approved for emergency use in the European Union [5], 121 candidate vaccines in clinical evaluation and 194 in preclinical studies [6], with more vaccines on track to complete clinical trials and possibly receive regulatory authorization for use.

To evaluate a vaccine candidate’s efficacy and safety, it is critical that vaccine trials are conducted in the right populations at the right time to improve the power of an accurate result for the product’s true efficacy, and to reduce the required sample size. Ideally, randomized controlled trials for vaccines are conducted in sites and population groups where there is a high incidence rate and where a significant proportion of the population remains susceptible to infection. Infectious disease models can help support the decision-making process in vaccine trial site selection by predicting which settings and populations are most likely to have high incidence rates during periods when trials are scheduled to occur. In addition, models can assist in predicting the expected rate of infection endpoints that is needed to calculate the required study sample size. Between June and August 2020, we used an agent-based model, Covasim [7], to project COVID-19 incidence and diagnosis rates for 72 locations across Australia (AU), Belgium (BE), Brazil (BR), France (FR), Italy (IT), Mexico (MX), the Netherlands (NL), South Africa (ZA), Spain (ES), and the United States (US). The Covasim model was used as one of multiple models to predict the incidence rate of infections and diagnosed (i.e., tested positive) cases for the 6-week window of 15 September to 31 October 2020, which was 2-3 months in the future from the time that the model projections were created. The 6-week window was the time during which the Janssen COVID-19 vaccine was scheduled to start enrolment in efficacy clinical trials. Model projections from multiple models (e.g., the MIT model), including the Covasim model, were considered in the selection of clinical trial sites alongside logistic, feasibility, time to enrolment, and other factors in vaccine trial site selection.

Infectious disease models, no matter how sophisticated, are inherently limited by the data used to calibrate them and the assumptions they make about the future conditions. Model validation is an important mechanism to understand which inputs and assumptions play the greatest role in determining model accuracy. For the 72 settings modelled, the projection period has now passed, meaning that it is possible to compare model outcomes with actual outcomes in each setting and to determine any correlates of accuracy. This information is valuable for improving future reliability of infectious disease models, as well as for improving our fundamental understanding of the COVID-19 pandemic.

In this study, we assess whether the actual data for each of the 72 settings fell within predicted confidence limits, the a priori accuracy of the ranking of COVID-19 projected incidence for potential trial sites, and usefulness of a statistical regression model to identify policy, socioeconomic, and other factors as predictors of model accuracy.

## 2. Methods

### 2.1 Model overview

Covasim is an agent-based microsimulation model that allows assessment of COVID-19 epidemic trajectories. The model can be calibrated to a given setting using available demographic data (population size, population age structure, household size distribution), setting-specific contact data (separated by household, school, work, and community contacts [8]), and global COVID-19 disease parameter estimates. It allows interventions including testing, contact tracing, quarantine, mask use, and social restriction policies (e.g., lockdowns, border controls, physical distancing) to be implemented with different levels of effectiveness and adherence. Once calibrated to match historical epidemiological data from the application context (e.g., number of tests, number of positive diagnoses, number of COVID-related deaths) and data or assumptions on transmission-related behaviors in the population and associated policies, the model can project the likely timing, duration and size of epidemic trajectories, including subsequent epidemic waves, based on input assumptions for future testing rates and policies. A detailed description of the Covasim methodology can be found in [7], with relevant applications in [9-11]. The models were generated with a population of 100,000 agents representing individuals who interact over common social layers (household, school, work, and community networks), and Covasim applies a dynamic scaling factor to the results to quantify the extent of the epidemic with respect to the size of the population in each setting.

### 2.2 Data and inputs

The modelling was conducted for 72 nominated settings (51 cities, 5 US counties, 6 French departments, 5 South African provinces, 3 Italian provinces, and 2 Australian states). Of the 51 cities, 3 were in Belgium, 13 in Brazil, 4 in Italy, 2 in Mexico, 2 in the Netherlands, 2 in Spain, and 25 in the US.

Time-varying data on COVID-19 testing, cases, and deaths were obtained from publicly-available government, state, and territory health department sources. In some instances, it proved difficult to source authoritative data detailed at the city level and, in such instances, data from broader areas such as counties, departments, and provinces were used to estimate city-level values by factoring in the population ratio.

### 2.3 Patient and Public Involvement

Study participants or the public were not involved in the design, or conduct, or reporting, or dissemination plans of our research.

### 2.4 Scenarios projected

At the time model scenarios were run (during July and August 2020), it was unclear what policy pathways each setting would take as they emerged from initial lockdowns; however, we assumed that it was unlikely to be a sudden policy change with return to pre-COVID behaviors in one step. So, for each location, we simulated five scenarios based on plausible future dates and intensities for policy changes that may occur, with the lower and upper bounds across scenarios used as an uncertainty interval for projected outcomes. The scenarios are defined as follows:

- **No changes to restrictions**: policies that were implemented at the data end date remained in place for the duration of the simulation,
- **10% easing of restrictions**: a 10% increase in average individual-level transmission, implemented (a) 6- or (b) 10-weeks from the data end date, to simulate a decrease in the efficacy of the policies in place / an easing of restrictions following a lockdown, and
- **20% easing of restrictions**: a 20% increase in average individual-level transmission, implemented (a) 6- or (b) 10-weeks from the data end date, to simulate a further decrease in the efficacy of the policies in place / an easing of restrictions following a lockdown.

Additionally, for the duration of the projection period, models were specified as having zero imported infections and a constant number of daily tests equal to the 7-day rolling average at the end of the available data. For settings that were not in lockdown at the time of the analysis but who were experiencing a resurgence of COVID-19 cases, analogous scenarios with increased restrictions were modelled.

### 2.5 Calibration to existing data

Calibration of the models was performed to fit the reported COVID-19 diagnoses and deaths over time in each setting by adjusting the overall transmission probability, the effectiveness of the past policy changes and the proportion of people with COVID-19 symptoms being tested. Where the mortality data appeared inconsistent with diagnoses data, given testing rates, the mortality data was judged to be the more reliable indicator. For each setting, past lockdown or policy changes were implemented in the models on the dates that they occurred, with the effectiveness of each lockdown or policy change deduced by calibrating the impact parameter in the model such that models reproduced the best fit to reported diagnoses and deaths. Simulations were run from the start date of the data for a given setting to October 31, 2020.

### 2.6 Outcome measures

The primary outcomes of each setting from September 15 to October 31, 2020 were the projected total number of new infections, the average daily incidence rate and the average daily diagnosis rate over a 30-day period, as well as the projected seroprevalence at the end of October. Upper and lower bounds for outcomes were based on the maximum/minimum values of the medians from the scenarios described in Section 2.3.

To evaluate the accuracy of projections, we assessed whether the actual data for number of new diagnoses for the period September 15 to October 31 was within the upper and lower bounds that were projected.

### 2.7 Statistical regression to determine correlates of accurate projections

Post hoc, we hypothesized that covariates associated with the modelled context might explain why our previous model projections were either consistent or inconsistent with the observed epidemic trajectory. We used a logistic regression model with 17 modelling, socioeconomic, demographic, health, and climate variables against the dependent variable of whether the observed data was within or outside the projected incidence range for modelled settings. Modelling variables included future policy assumption accuracy, defined as being correct if actual policy changes (easing or increasing of restrictions) occurred within a month of the dates they were modelled; future testing assumption accuracy, defined as being correct if modelled testing rates fell within a 25% margin of the actual ones; future border control assumption accuracy, defined as being correct if borders remained closed as we had assumed in our models; days between the end of the data (i.e. calibration end date) and September 15, 2020; and two parameters assessing calibration accuracy, the average difference between the rate of change of reported cumulative diagnosis and deaths versus these modelled indicators over the calibration period (the difference in rate of change was selected over just the observations to capture a stricter calibration criteria). The final multivariate regression only considered variables that were statistically significantly correlated with the outcome variable in the univariate analyses (p < 0.1 was chosen here). A sensitivity analysis was tested where a random-effects term was included for country to determine whether it might be influencing the outputs.

## 3. Results

### 3.1 Illustrative examples of original projections: Paris (FR) and New York (US)

Calibrations, projections, and summary model outcomes are presented for the major cities of Paris (population of 2.15 million) and New York (population of 8.34 million) in **Figures 1 and 2** and in **Table 1**, respectively. The calibration period corresponding to the available data at the time of study was from February 25 to August 24, 2020 in Paris, and from February 20 to July 7, 2020 in New York. Paris was an example of a city where the observed data did not fall within the upper and lower bound of the projections. In contrast, New York is an example of a city where the observed data was within the upper and lower bounds of the scenarios.

**Figure 1:**
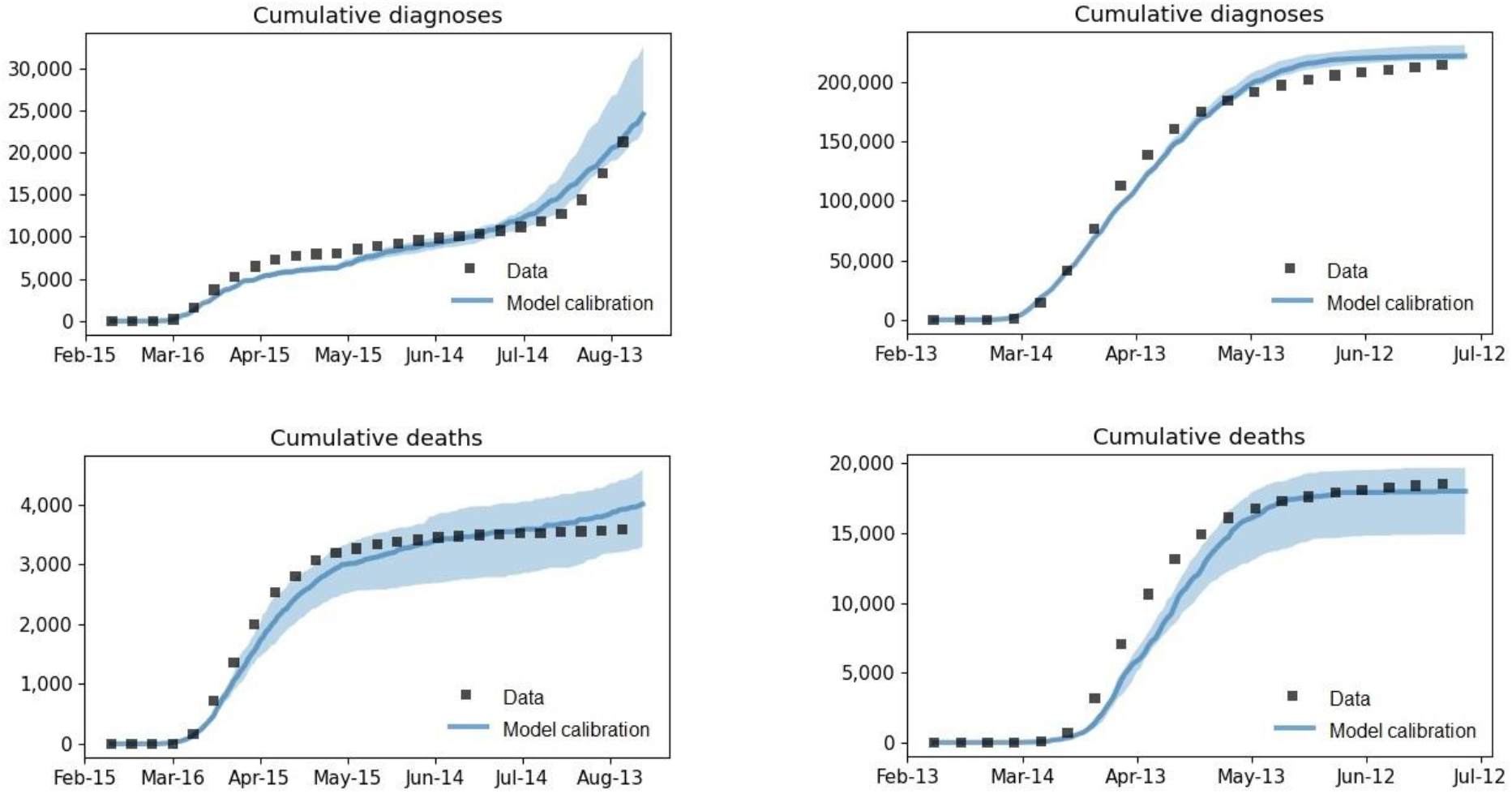
calibration of Paris (left) and New York (right). Black dots represent the data, the blue line the median model projection, and the blue shaded area the confidence interval.

**Figure 2:**
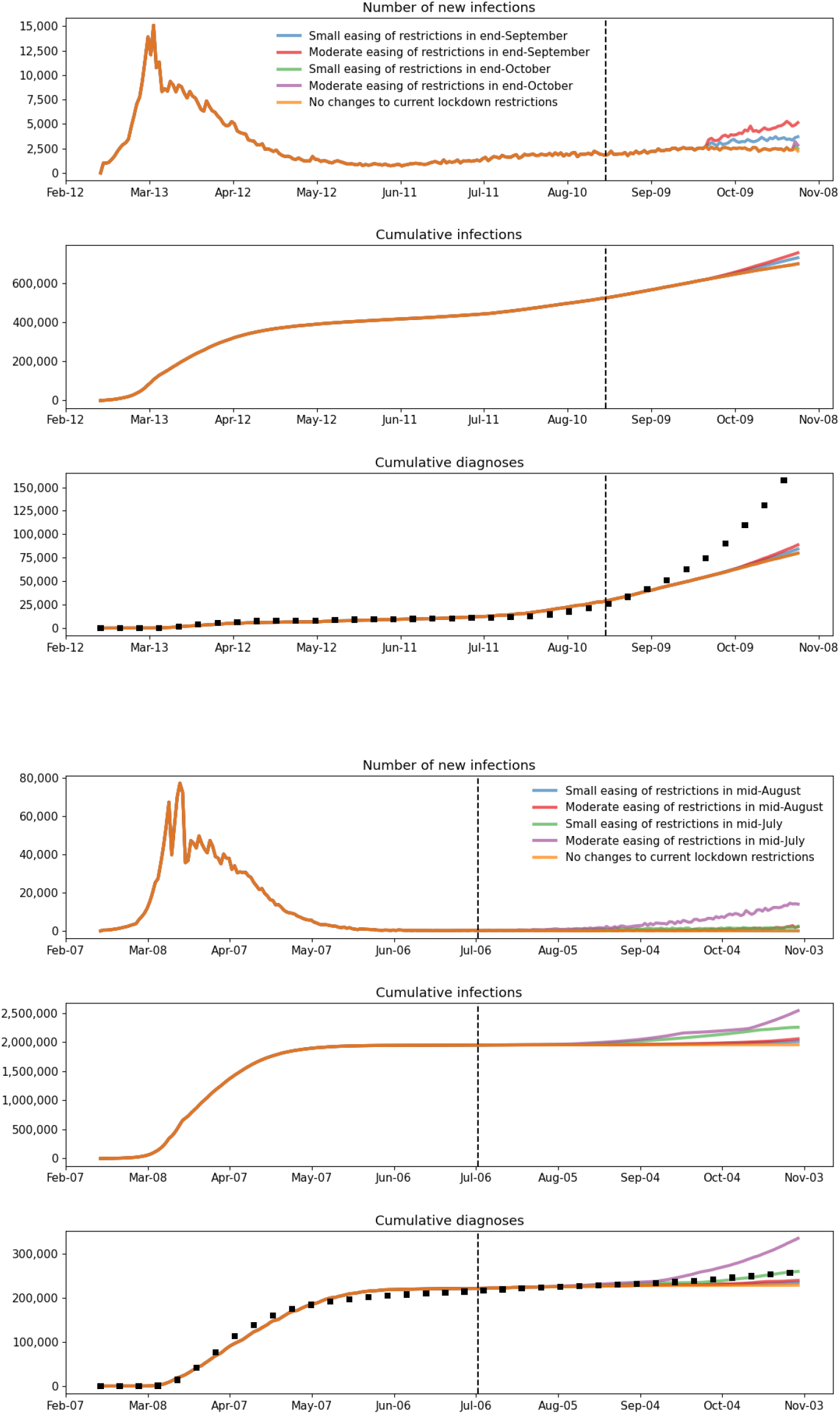
Number of new infections, cumulative infections, and cumulative diagnoses in Paris (top) and New York (bottom). The dashed vertical line represents the start of the model projection period, the black squares the actual data and the colored lines the scenarios modelled.

**Table 1:** Model-projected COVID-19 outcomes and actual reported data for the cities of Paris and New York from September 15 to October 31, 2020.

|  | Model: number of new SARS-CoV-2 infections |  | Model: 30-day SARS-CoV-2 infection incidence rate* |  | Model Predicted: seroprevalence end of October |  | Model predicted and actual data: 30-day detected cases (/population size) |  | Data |
| --- | --- | --- | --- | --- | --- | --- | --- | --- | --- |
|  | Lower Bound** | Upper Bound** | Lower Bound** | Upper Bound** | Lower Bound** | Upper Bound** | Lower Bound** | Upper Bound** |  |
| **Paris** | 51,169 | 82,006 | 1.55% | 2.49% | 0.27% | 0.28% | 0.41% | 0.52% | 3.70% |
| **New York** | 741 | 416,358 | 0.01% | 3.26% | 0.22% | 0.25% | 0.00% | 0.64% | 0.18% |

\* New SARS-CoV-2 infections occurring between September 15 and October 31, 2020 were converted to a 30-day incidence rate for comparability
\*\* The lower and upper bounds were taken as the minimum and maximum median values, respectively, of the five forecasting scenarios

### 3.2 Assessment of outcomes: detected cases

Of the 72 locations modelled, 27 (37.5%) yielded correct projections (observed data was within projected range) and 45 (62.5%) were out of range. The 30-day case detection rate from the projections and the data are illustrated in **Figure 3**. Epidemic trajectories for detected cases were under-estimated in 78% of settings with incorrect projections.

**Figure 3:**
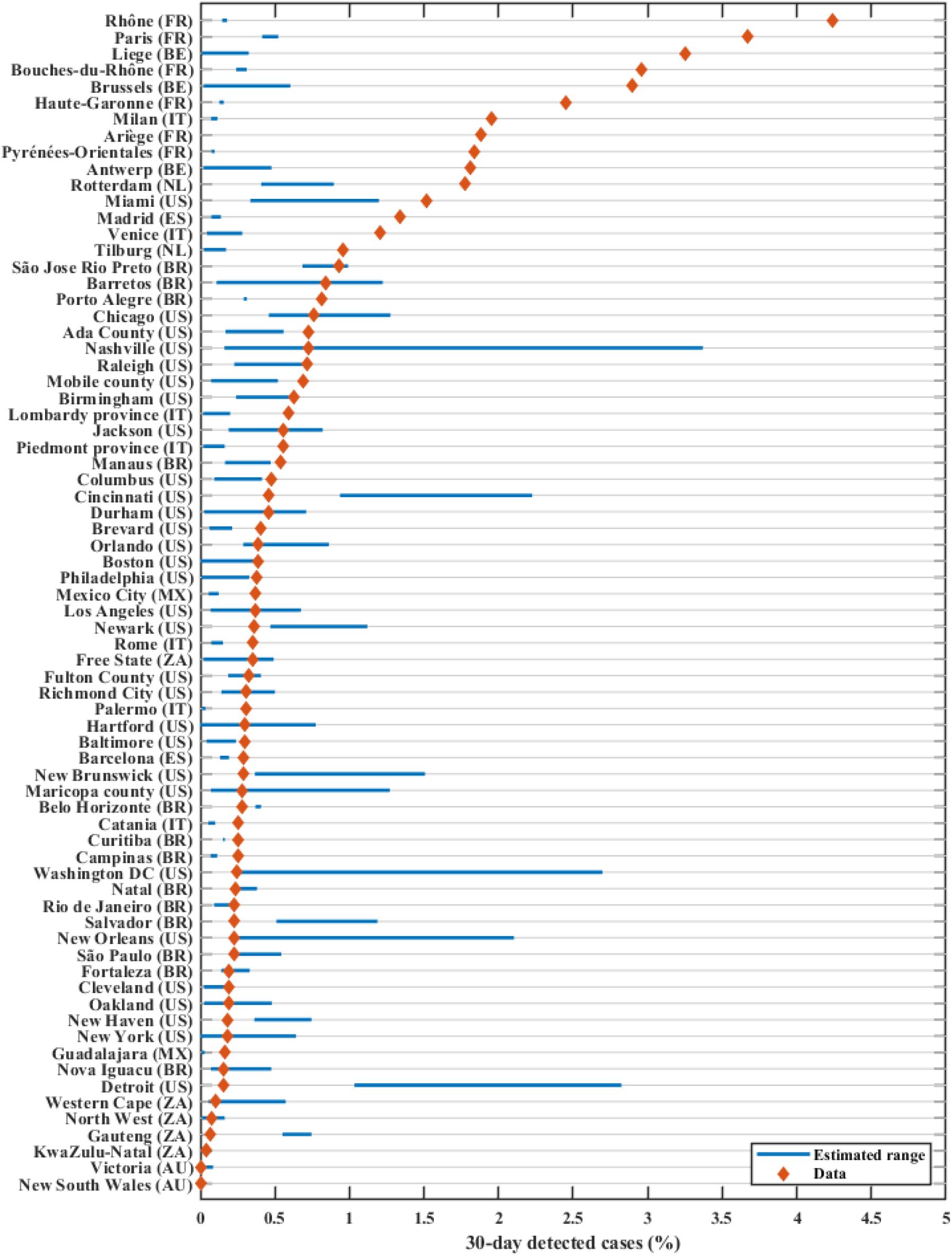
30-day detected COVID-19 cases for September 15 to October 31, 2020, sorted by data in descending order. The blue line represents the estimated range from the simulations, and the red diamond the data.

### 3.3 Logistic regression

Univariate analysis showed that correct future policy assumptions, correct testing rates, longer projection periods, a higher poverty rate, a higher unemployment rate, a larger proportion of the population younger than 30 and a smaller proportion of the population older than 65 were correlated with correct model projections (**Table 2**). Multivariate logistic regression revealed that, by far, the most important predictor for accurately predicting the epidemic trajectory as compared with observed data is the correctness of assumed future policy changes (or scenarios) implemented in the models (p < 0.05). A sensitivity analysis indicated that this was true even when a random-effects term was included for country.

**Table 2:**
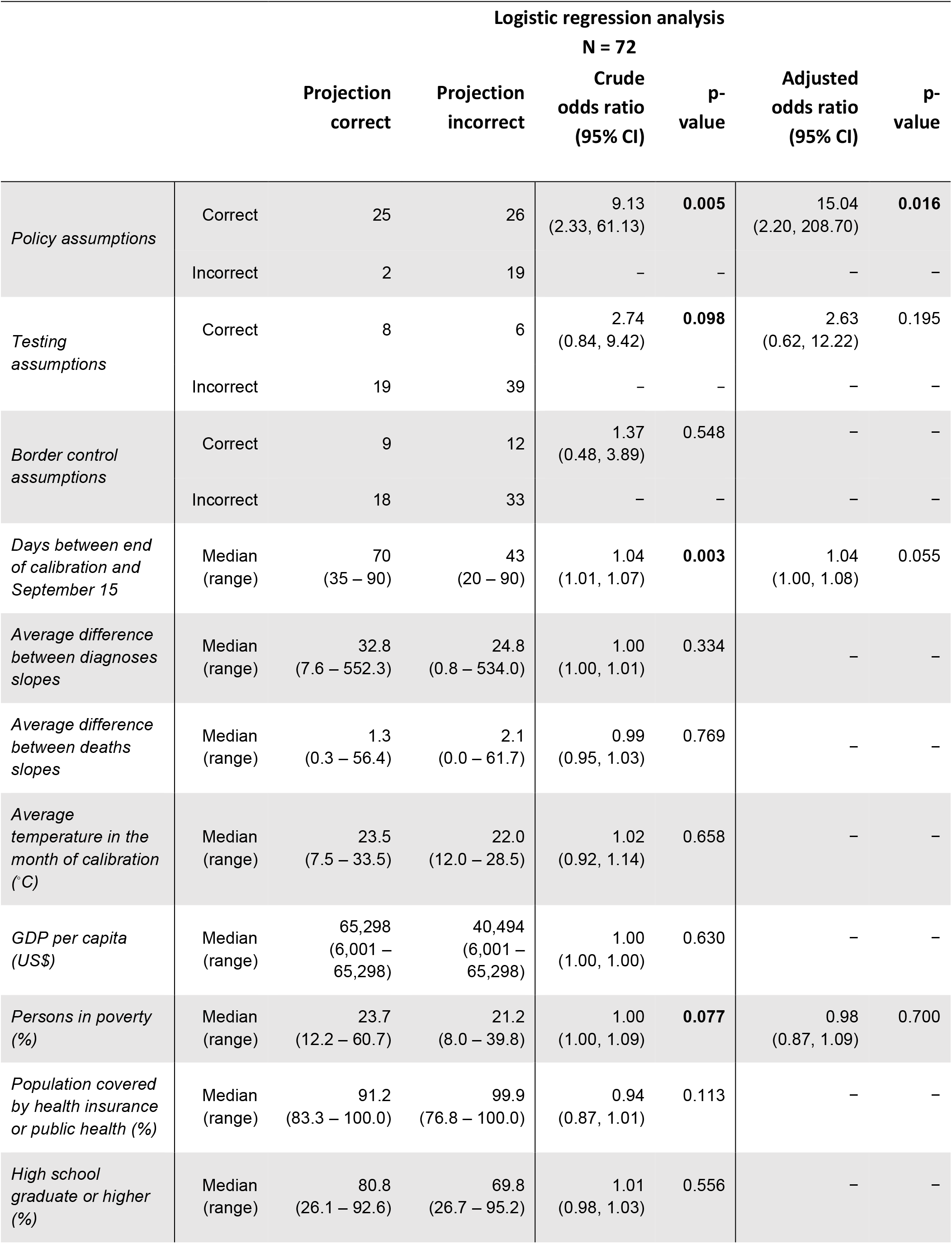
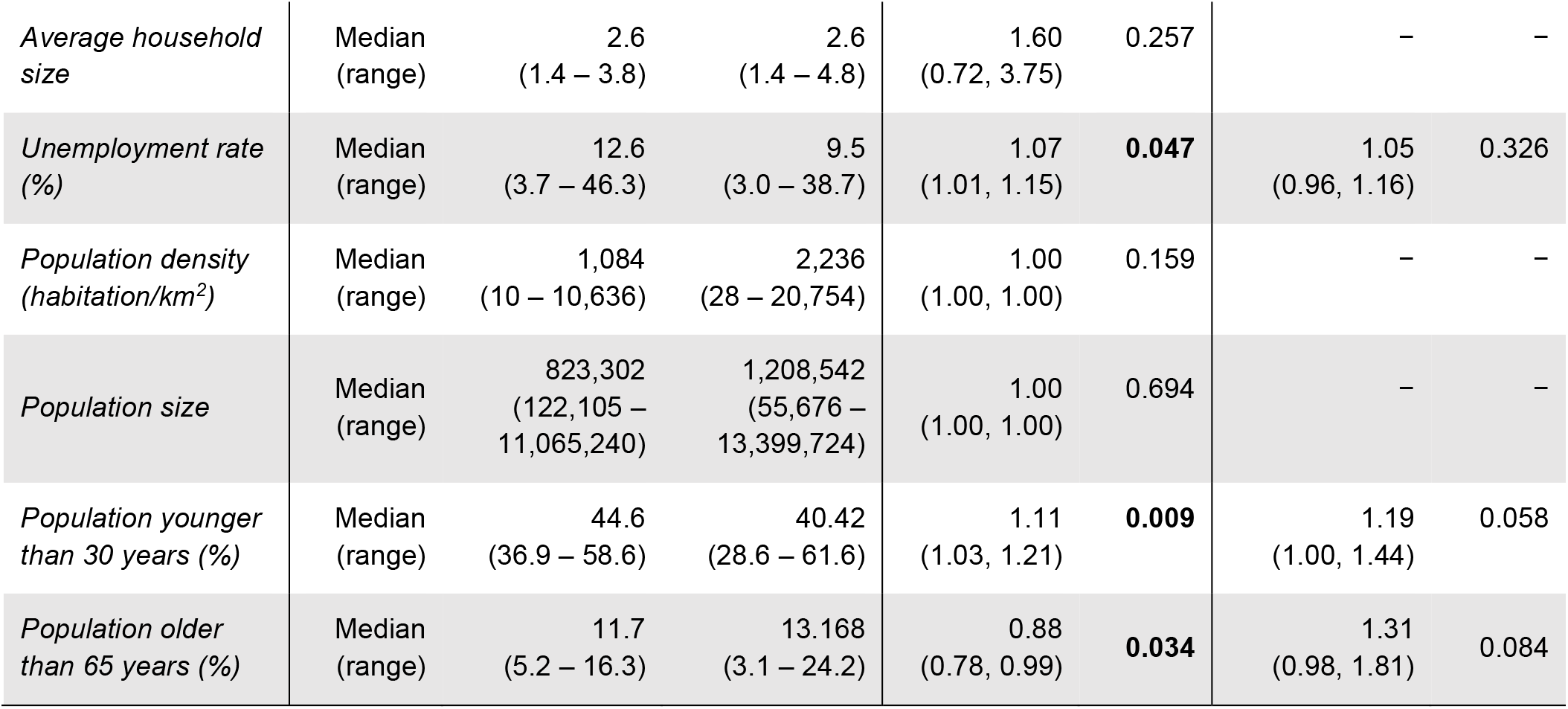
Logistic regression results for model projection accuracy.

## 4. Discussion

Proper model validation is an important yet often overlooked aspect of modelling. With the growing use of mathematical modelling to inform and guide major decisions being made in public health, validating past model projections against real-world data can provide valuable insight into how models can be improved and fine-tuned for future studies [12]. Eaton et al.’s paper on human immunodeficiency virus (HIV) modelling [13] is an example of such work: data from the 2012 household survey in South Africa was used to evaluate the accuracy of ten previous model projections of HIV prevalence and treatment coverage in 2012, with findings emphasizing that model projections consistently under-predicted HIV incidence and prevalence for that year. More recently, James et al. [14] wrote of the key limitations of mathematical modelling as a tool for policymaking in the context of the global health crisis wrought by COVID-19. More specifically, describing how the rapidly changing epidemiological situation coupled with the fact that the effectiveness of new policies may not be well documented poses challenges in the evaluation and implementation of new policies in COVID-19 models. Indeed, since the start of the pandemic, COVID-19 predictions have differed significantly between the different epidemiological models and have received criticism for being wrong [15, 16]. For instance, in the initial stages of the epidemic in the United States, the number of deaths from COVID-19 on any single day fell outside the predicted range of some prominent models up to 70% of the time [17]. Eleven models assembled by FiveThirtyEight show how estimates can vary widely due to the different underlying assumptions made about policy changes and the number of contacts in the models [18]. The Centers for Disease Control and Prevention in the United States has compiled a list of 37 independent models in its published weekly forecasts predicting national and state numbers of new COVID-19 cases, deaths, and hospitalizations in the United States [19]. Moreover, these model outputs were compared against each other, validated against empirical data, and aggregated into an “ensemble” forecast to improve the predictive performance.

In this study we revisited COVID-19 epidemic projections for 72 candidate clinical trial settings to identify policy, socioeconomic, and other factors that could be predictors of model accuracy. Of the 17 covariates evaluated, we found that making correct assumptions about future policies and restrictions was the most significant driver of whether projections were accurate or not. These results, combined with the frequent changes in restrictions implemented by governments throughout the pandemic to date (at the time of the analysis), help to explain why past model projections were inaccurate. These findings also support the need for frequent revision of projections as policies change and highlight the risks of using a model to project COVID-19 epidemics too far into the future—although how far remains unclear.

A limitation to this study, and the original modelling, is that it can be difficult to predict the intensity and timing of future policy changes, let alone translate real-world policy changes and restrictions into reductions in transmission risk in the model. For this study we classified our policy change assumptions as being incorrect when the actual restriction change was actually made in the opposite direction to our assumption (e.g., we modelled a tightening of restrictions and an easing of restriction occurred) or when the timing of the policy change occurred more than a month from when we modelled it to occur. It is not surprising that these scenarios led to out-of-range projections: the projection for a scenario is not inaccurate if the scenario never happened. However, in instances where the actual policy change occurred at the time that we assumed, forecast accuracy was only 49%. This could have occurred because either behavioral or health systems changes occurred that could not be quantified and captured in the model, or because the policy and restriction changes had more or less impact than we assumed. For policy changes occurring in the past, the effect size of the restrictions can be calibrated; however, for future policy changes this is not possible, and the effectiveness of future restrictions must be estimated based on the impacts they have had in the past. This introduces uncertainty because people’s behavior in response to public health directions may not be consistent over time.

There are other limitations to COVID-19 modelling that may explain discrepancies in model projections. First, diagnoses and deaths are often underreported due asymptomatic cases, limited testing, limited health system capacity, or limited (or suppressed) reporting. Second, transmission is highly heterogeneous and even data at a city-level may result in epidemic behavior being smoothed out. Third, some of the data used to calibrate the models (up to July 2020) is likely to be outdated due to the emergence of new COVID-19 variants with increased transmissibility and mortality [20]. One example that is relevant to this study is in Brazil, where the SARS-CoV-2 variant P.1 emerged as early as July 2020 and is estimated to be 1.4 to 2.4 times more transmissible than previously circulating variants [21-23] (in Brazil, the epidemiological data was within the projected range 46% of the time). Fourth, with more than 5.7 billion vaccine doses administered globally as of September 22, 2021 [1], vaccine roll-out plays a significant role in transmission from 2021. In the context of today’s highly transmissible Delta variant, the impact of the different vaccines and the waning immunity on severe disease and deaths must be modelled accordingly, as well as (from a policy perspective) the deployment of booster doses including which vaccines will be used, when, and for which target populations.

## 5. Conclusions

Our findings suggest that the policies administered in response to a pandemic are of utmost importance when it comes to determining epidemic outcomes. However, the frequent, somewhat unpredictable, and significant changes in restrictions implemented heterogeneously across settings can largely explain why COVID-19 projections may be inaccurate. Therefore, epidemic modelling for informing epidemic mitigation should be conducted in unison with policy makers and be driven by the needs of these decision makers, and epidemic modelling for research and trial purposes should be updated regularly with the best possible data, assumptions and sensitivity analyses, and should involve the collaborative insight of behavioral and policy experts.

## Data Availability

Data are available upon request.

## Contributors

NS, PC and DPW conceived and designed the original modelling study. SWH, RA, RMS and CCK developed the Covasim model. FH, AP, DD, TW and SLK conducted the original data collection and modelling. FH conducted the post hoc data collection, post-processing and statistical analyses. FH prepared the write-up and NS supervised the writing and editing of the manuscript. All authors have read, contributed to and approved of the final version of the paper. NS is the guarantor and joint senior author of the manuscript.

## References

1 World Health Organization. WHO Coronavirus (COVID-19) Dashboard. [cited 2021 September]; Available from: https://covid19.who.int/.

2 World Health Organization. In WHO global pulse survey, 90% of countries report disruptions to essential health services since COVID-19 pandemic. 2020 August 31; Available from: https://www.who.int/newsroom/detail/31-08-2020-in-who-global-pulse-survey-90-of-countries-report-disruptions-to-essential-healthservices-since-covid-19-pandemic.

3 The World Bank. 2020 Year in Review: The impact of COVID-19 in 12 charts. 2020 [accessed 2021 June]; Available from: https://blogs.worldbank.org/voices/2020-year-review-impact-covid-19-12-charts.

4 The World Bank. COVID-19 to Add as Many as 150 Million Extreme Poor by 2021. 2020 [cited 2021 June]; Available from: https://www.worldbank.org/en/news/press-release/2020/10/07/covid-19-to-add-as-many-as150-million-extreme-poor-by-2021.

5 Zimmer C, Corum J, Wee S-L. Coronavirus Vaccine Tracker. Available from: https://www.nytimes.com/interactive/2020/science/coronavirus-vaccine-tracker.html.

6 World Health Organization. Draft landscape of COVID-19 candidate vaccines. 2020 September 8; Available from: https://www.who.int/publications/m/item/draft-landscape-of-covid-19-candidate-vaccines.

7 Kerr CC, Stuart R M, Mistry D, et al. Covasim: an agent-based model of COVID-19 dynamics and interventions. PLoS Comput. Biol.2021;17(7):e1009149.

8 Prem K, Cook AR, Jit M. Projecting social contact matrices in 152 countries using contact surveys and demographic data. PLoS Comput. Biol.2017;13(9):1–21.

9 Kerr CC, Mistry D, Stuart R M, et al. Controlling COVID-19 via test-trace-quarantine. Nat Commun.2021;12(1):2993.

10 Panovska-Griffiths J, Kerr CC, Stuart R M et al. Determining the optimal strategy for reopening schools, the impact of test and trace interventions, and the risk of occurrence of a second COVID-19 epidemic wave in the UK: a modelling study. Lancet Child Adolesc. Health2020;4:817–27.

11 Scott N, Palmer A, Delport D, et al. Modelling the impact of reducing control measures on the COVID-19 pandemic in a low transmission setting. Med J Aust2021;214(2):79–83.

12 Wilson DP and CC Kerr. Can we know in advance whether models will get it right? Lancet Glob. Health2015;3(10):E577–E578.

13 Eaton JW, Bacaër N, Bershteyn A, et al. Assessment of epidemic projections using recent HIV survey data in South Africa: a validation analysis of ten mathematical models of HIV epidemiology in the antiretroviral therapy era. Lancet Glob. Health2015;3(10):e598–e608.

14 James LP, Salomon JA, Buckee CO, et al. The Use and Misuse of Mathematical Modeling for Infectious Disease Policymaking: Lessons for the COVID-19 Pandemic. Med Decis Making2021;41(4):379–385.

15 Holmdahl, I and Buckee C. Wrong but Useful — What Covid-19 Epidemiologic Models Can and Cannot Tell Us. N Engl J Med 2020; 383:303–305.

16 Kreps SE, Kriner DL. Model uncertainty, political contestation, and public trust in science: Evidence from the COVID-19 pandemic. Sci. Adv.2020;6(43).

17 Marchant R, Samia NI, Rosen O, et al. Learning as We Go: An Examination of the Statistical Accuracy of COVID19 Daily Death Count Predictions. 2004.04734 [stat.OT] 2020.

18 Best R, Boice J. Where the latest COVID-19 models think we’re headed—and why they disagree. 2020 [accessed 2021 May]; Available from: https://projects.fivethirtyeight.com/covid-forecasts/.

19 Centers for Disease Control and Prevention. COVID-19 Forecasting: Background Information. 2020 [accessed 2021 May]; Available from: https://www.cdc.gov/coronavirus/2019-ncov/casesupdates/forecasting.html.

20 Centers for Disease Control and Prevention. SARS-CoV-2 Variant Classifications and Definitions. [cited 2021 June]; Available from: https://www.cdc.gov/coronavirus/2019-ncov/variants/variant-info.html.

21 Voloch CM, da Silva F Jr R, de Almeida Luiz GP, et al. Genomic characterization of a novel SARS-CoV-2 lineage from Rio de Janeiro, Brazil. J Virol2021;95(10):e00119–21.

22 Faria NR, Mellan TA, Whittaker C, et al. Genomics and epidemiology of the P.1 SARS-CoV-2 lineage in Manaus, Brazil. Science2021;372(6544):815–821.

23 Coutinho RM, Marquitti FMD, Ferreira LS, et al. Model-based evaluation of transmissibility and reinfection for the P.1 variant of the SARS-CoV-2. medRxiv2021.03.03.21252706

